# Risk stratification in hip and knee replacement using Artificial Intelligence: a dual centre study to support the utility of high-volume low-complexity hubs and ambulatory surgery centres

**DOI:** 10.1101/2024.11.29.24317637

**Authors:** Christopher Woodward, Justin Green, MR Reed, David J Beard, Paul R Williams

## Abstract

The COVID-19 pandemic has resulted in a significant backlog of hip and knee replacement surgeries in the United Kingdom (UK). ^1,2^ To address this, surgical hubs have been proposed to enhance efficiency, particularly for high-volume, low-complexity cases. ^3,4^ These hubs and Ambulatory Surgery Centres often lack higher level care support such as intensive care facilities and are thus suited to patients with less co-morbidity and systemic illness. Pre-operative risk assessment is required to enable correct patient allocation to the appropriate site and reduce unwarranted risk.

This study explores the use of artificial intelligence (AI) for risk stratification in hip and knee arthroplasty. A polynomial regression model was developed using patient demographics, blood results, and comorbidities to assign risk scores for postoperative complications. The model was generated from 29,658 patient records from two UK National Health Service (NHS) healthcare organisations. It demonstrated an area under the receiver operating characteristic curve (AUROC) as the evaluation metric and was capable of categorising patients into high and low risk. Validation was performed using a retrospective analysis of 445 patients. Predicted versus actual complications and need for further care were used to examine agreement. The model’s sensitivity was 70% for identifying high-risk patients and had a negative predictive value of 96%. This AI risk prediction was comparable to consultant-led care in risk stratification.

These findings suggest that AI can support more streamlined and efficient preoperative risk stratification, potentially reducing the burden on preoperative assessment teams and optimising resource allocation. While not without limitations, the AI model offers a sophisticated adjunct to clinical decision-making around determining risk. This can support facilities like hubs in the UK NHS or Ambulatory Surgery Centres in the United States.

## Background

In the United Kingdom there remains a significant backlog of hip and knee replacement surgery following the COVID 19 pandemic.^1,2,3^ The proposed rapid significant and sustained expansion in services required to address the deficit may be best addressed in surgical hubs.^4,5^ Compared to standard NHS systems, hubs offer increased productivity equating to 11% extra cases for established centres and 22% new centres. A hub is defined as a ring-fenced facility which exclusively delivers elective care and given that they often lack intensive care or high dependency facilities, have a necessary focus on high-volume-low complexity cases. These institutions can provide surgical treatment for patients with lower complexity co-morbidities, and in doing so can optimise throughput.

Other hospitals with facilities to address patients with more complex needs such as Intensive Care Units are often termed Low Volume High Complexity (LVHC) sites.^6^ In the United States, due to economic issues and rule changes, the use of Ambulatory Surgery Centres (ACS) has increased dramatically in recent years with an 84% increase in activity in 2022 alone.

Both hubs and ambulatory surgery centres face similar issues with respect to optimising patient selection. Emergency transfer of a patient who develops an unforeseen complication in a hub or ACS is both dangerous and expensive. Appropriate patient allocation to these sites is a key feature for success, and pre-operative assessment of risk and health maximisation, poses a burden within in preoperative assessment teams. It is postulated that Artificial Intelligence (AI) can assist with this problem.^7^

The use of artificial intelligence is increasing in medicine and surgery, with several uses suggested in orthopaedics, including risk prediction. ^7,8^ Other risk prediction tools have been used in surgery ^9,10^ but AI and machine learning offers a more sophisticated method,^11^ and has been trialled successfully in other specialties .^12,13^

Machine learning (ML), using varied comorbidities, demographics, and socioeconomic factors, has been employed to produce electronic risk calculators for patient counselling and predicting surgical outcomes.^14^ Recently, ML algorithms have gained popularity for capturing complex non-linear relationships within data. ^15^ ML prediction models are useful for setting patient expectations, preparing for outcomes, and managing complications.^16^ Comparative technology based on the P-POSSUM risk stratification model accurately predicted morbidity and mortality following hip surgery. ^17,18^ However, small sample sizes and study ambiguity provided inconclusive results.

In part 1 of the work, we use a polynomial regression prediction model, that incorporates patient demographics, blood results, and comorbidities from two sources, to assign each individual patient a percentage risk score of the likelihood of developing a post-operative complication. The intention is to help stratify pre assessment requirements, as well as informing site-specific theatre list and hospital site capacity. It could aid in preoperative patient optimisation to mitigate risks, thereby improving patient safety. In addition, patients can understand their own risk to make informed decisions.

In part 2 of the work, we perform a validation study in which retrospective hip and knee arthroplasty data from two UK healthcare organisations, Swansea Bay Health Board and Northumbria Healthcare NHS Foundation Trust, is used to test the accuracy of the AI prediction. The intention was to identify how many patients were correctly assigned the category of “high risk” by the AI model. Sensitivity and Negative Predictive Value (NPV) analysis are used as indices for accuracy.

Overall, the aim was to assess the feasibility of using the model, as a more streamlined and efficient method, to prospectively predict risk in a larger population (as part of the development sequence). Such a model could aid decisions made by perioperative teams.

## Methods

### Part 1: Model development

A dataset of 29,658 elective primary total joint replacement patient records from the two healthcare providers between January 2014 and March 2020, was analysed (Northumbria Healthcare NHS Foundation Trust n= 15,022, Swansea Bay University Health Board n=14,636).

There are 26 features (data points) used as predictors in the model, which are grouped into three categories: patient information, comorbidities, and blood measurements. 56.6% data points describe patients undergoing knee surgery and 43.4% hip surgery. Non-numeric entries such as those that might indicate laboratory values, were discarded and data converted to contain purely numeric data types, ensuring they can be used in computational models. The study included patients who underwent only primary total hip replacement procedures and primary total knee replacement procedures. Duplicate entries were deleted. A final dataset of n=9,137 records (237,562 data points) were used in the modelling.

A polynomial logistic regression machine learning model (AI) was developed in Northumbria Healthcare NHS Foundation Trust further refined in collaboration with SBUHB (Swansea Bay University Health Board). The model categorisation incorporated data from patient demographics, co-morbidities, blood tests, and overall health status (Table 1). Complications were considered for inclusion in the model (Table 2).

**Table 1:**
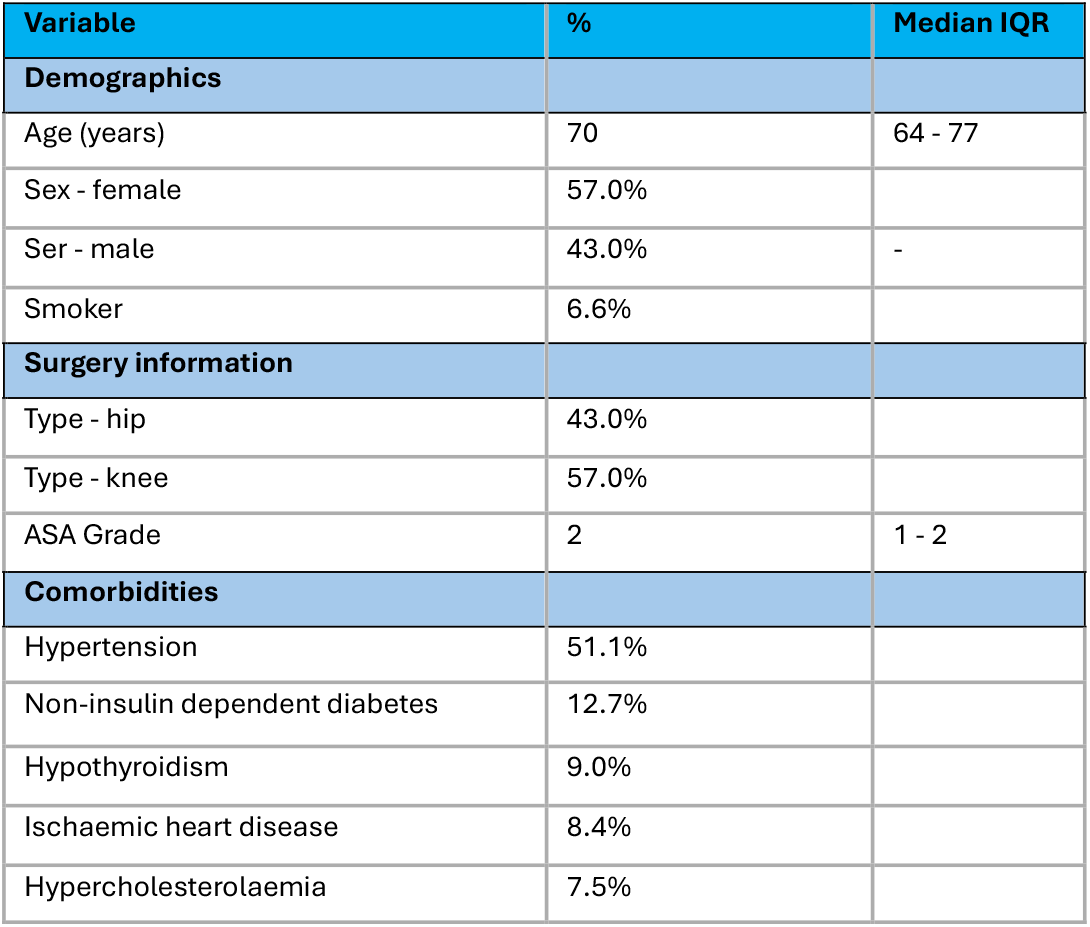
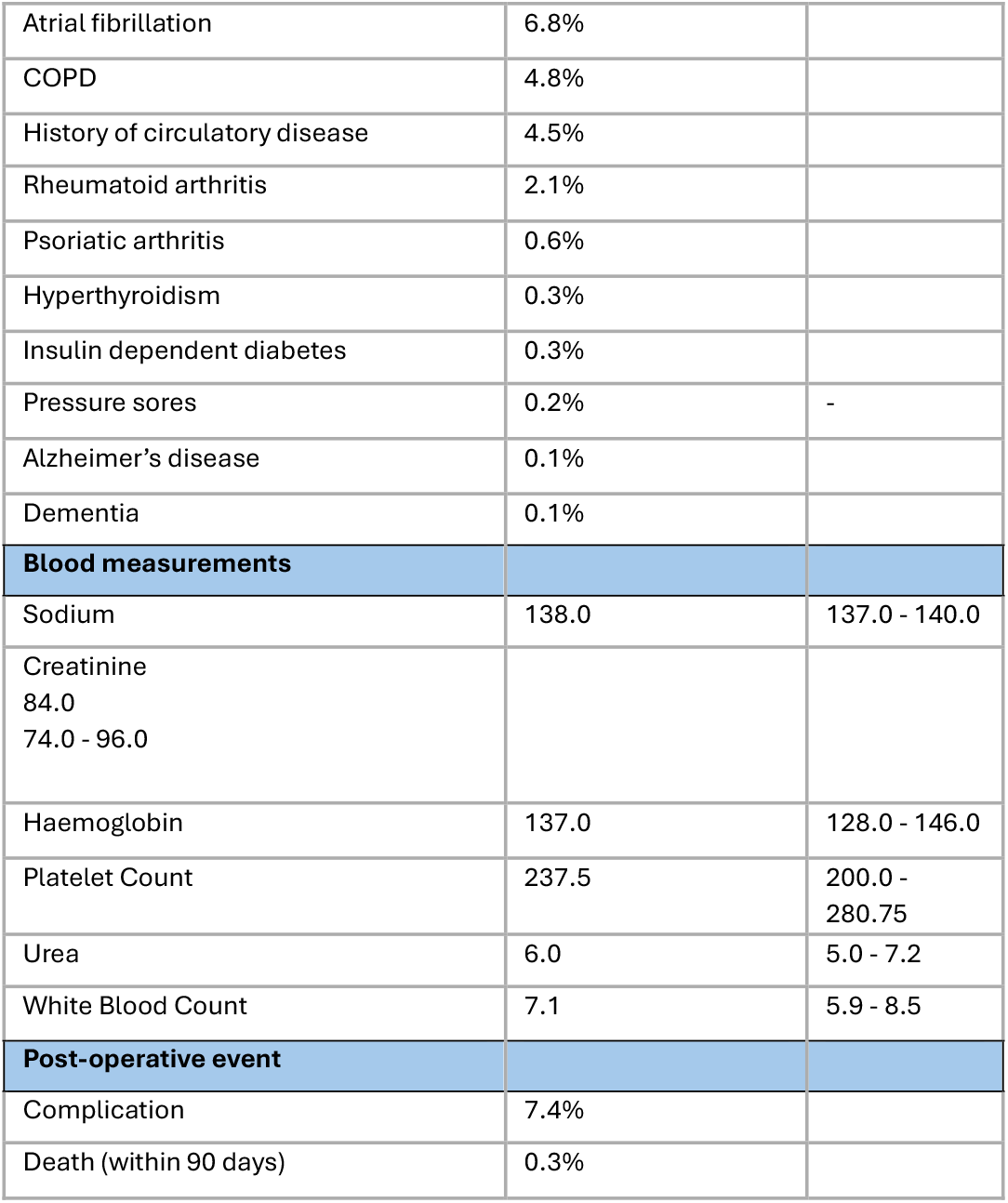
Data distribution of the population (n=6730). Variables with single asterisks are described by their median and interquartile range, otherwise in terms of their frequency.

| Variable | % | Median IQR |
| --- | --- | --- |
| **Demographics** |  |  |
| Age (years) | 70 | 64 - 77 |
| Sex - female | 57.0% |  |
| Sex - male | 43.0% | - |
| Smoker | 6.6% |  |
| **Surgery information** |  |  |
| Type - hip | 43.0% |  |
| Type - knee | 57.0% |  |
| ASA Grade | 2 | 1 - 2 |
| **Comorbidities** |  |  |
| Hypertension | 51.1% |  |
| Non-insulin dependent diabetes | 12.7% |  |
| Hypothyroidism | 9.0% |  |
| Ischaemic heart disease | 8.4% |  |
| Hypercholesterolaemia | 7.5% |  |
| Atrial fibrillation | 6.8% |  |
| COPD | 4.8% |  |
| History of circulatory disease | 4.5% |  |
| Rheumatoid arthritis | 2.1% |  |
| Psoriatic arthritis | 0.6% |  |
| Hyperthyroidism | 0.3% |  |
| Insulin dependent diabetes | 0.3% |  |
| Pressure sores | 0.2% | - |
| Alzheimer's disease | 0.1% |  |
| Dementia | 0.1% |  |
| **Blood measurements** |  |  |
| Sodium | 138.0 | 137.0 - 140.0 |
| Creatinine<br>84.0<br>74.0 - 96.0 |  |  |
| Haemoglobin | 137.0 | 128.0 - 146.0 |
| Platelet Count | 237.5 | 200.0 - 280.75 |
| Urea | 6.0 | 5.0 - 7.2 |
| White Blood Count | 7.1 | 5.9 - 8.5 |
| **Post-operative event** |  |  |
| Complication | 7.4% |  |
| Death (within 90 days) | 0.3% |  |

**Table 2:**
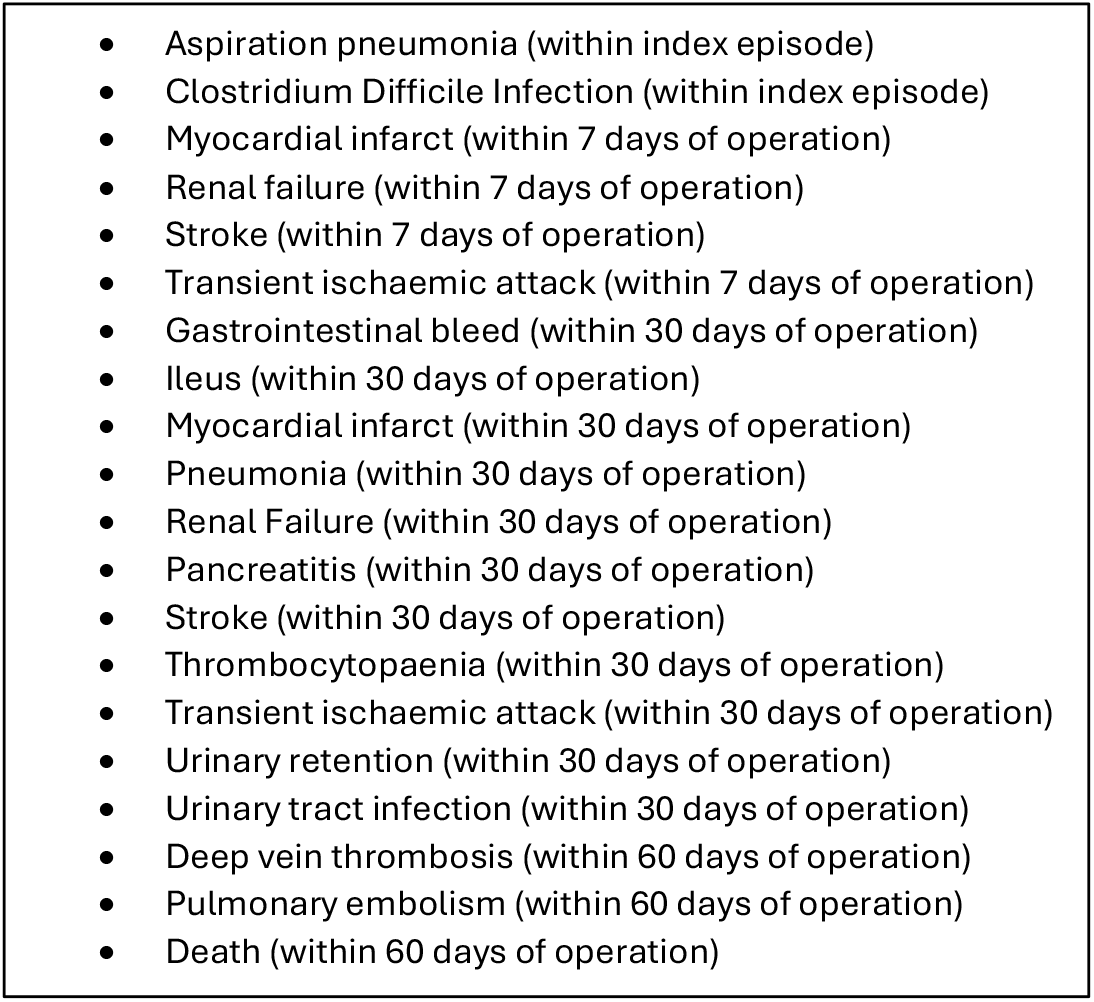
Medical complications considered as adverse outcomes in the modelling.

Information on each patient was curated based on previous evaluation of data relating to comorbidities, demographics, blood tests and contact with other specialties in an outpatient setting (Table 1).

Challenges in modelling include the risk of data leakage, a process where data is included in modelling that has decreasing relevance to the current practice. Factors, such as a change in healthcare providers delivery of service, e.g. consolidation of high complexity services into a specialist care centre and changes in clinical coding policy were identified, with the intention of ensuring relevance in modelling based on up-to-date practice.

To minimise this and ensure a realistic assessment of the model’s performance, the dataset was split along the time axis. The train set included patients who underwent surgery before August 2017 (n=6,730), while the test set consisted of patients who underwent surgery after this date. This resulted in a test set containing 18.5% of the total data points (n=43,949). It is important to note that some patients appeared more than once in the record having undergone multi joint procedures. To prevent data leakage, all patients who were included in the train set were removed from the test set. A test set of n=2,407 patient records were identified and divided based on type of operation, total knee replacement n= 1,035 (43%) and total hip replacement n=1,372 (57%).

The area under the receiver operating characteristic curve (AUROC) was used as the evaluation metric to choose the best combination of hyperparameters (**) for the model architecture. AUROC is a suitable choice for a risk stratification model, as it measures the model’s ability to differentiate between patients with and without complications, regardless of the chosen classification threshold.

To address the issue of imbalanced data, modelling was trained with and without oversampling in the training data. Oversampling is a commonly used technique for handling imbalanced datasets, as it creates a more balanced distribution of the target variable by replicating minority class instances. In our study, oversampling was implemented until a proportion of 50% was reached for each class, resulting in an optimally balanced training data set. This allows the model to better learn the characteristics of the minority class, potentially improving its performance in predicting postoperative complications.

Finally, the modelling was evaluated by the capacity to discriminate between output classes using AUROC (e.g. those encountering adverse events post operatively and those who didn’t). Second, the confidence in the predictions using the Brier score, which measures the mean squared difference between the predicted probabilities and the actual outcomes. Finally, the calibration of the predicted probabilities was assessed using the Expected Calibration Error (ECE), which measures the difference between the predicted and observed probabilities across various intervals.

Focussing on probabilities is essential in the case of a risk stratification model, as it allows the quantification of a patient’s risk of a complication. For ease of use the probability was converted into a categorical value that provided clinical users a grouping of patients. This, in turn, enables clinicians to identify high-risk patients who may require additional medical attention or closer monitoring, while not raising unnecessary alarms for low-risk patients. The thresholds were set as follows:

- Low risk of adverse outcome probability of 0.00 – 0.06
- Moderate risk of adverse outcome probability of 0.06 - 0.075
- High risk of adverse outcome probability of 0.075 and above

### Risk Ratio Thresholds

Thresholds to determine whether in a low risk or moderate to high-risk group was informed by a desire to achieve a meaningful reduction in relative risk from the baseline, consistent with the findings of previous studies that advocate for setting risk thresholds based on relative risk reductions to optimise patient management.^19,20^

Thresholds for risk for this study were determined by analysis of complications (Table 1), in the Northumbria cohort (143 patients). The complication rate in that unselected group was 7.5%. The using a probability of 0.06, i.e. 6% as a threshold was pragmatically based on a 20% relative risk reduction, from the overall baseline of 7.5% in that population, and being a useful cut off in terms of planning elective beds.

106 patients were deemed low risk i.e. 75%. From a clinical perspective, projecting to the planned number of cases undertaken at the each of the hospital organisations, this complemented the availability of around 1800 (of a possible 2400) elective operative sessions per year at the HVLC elective sites (hubs).

Patients with a less than 6% chance of complications were determined as low risk, with patients with a higher than 6% chance moderate to high risk.

### Part 2: Model evaluation and validation

Evaluation of the model was performed by retrospective analysis of 445 separate patient records in the two separate health organisations who had undergone elective primary hip and knee replacement surgeries using the ICD10 and OPCS4 frameworks of clinical coding.

The pre-operative electronic health records were digitally interrogated by the model to provide a risk stratification ratio percentage of developing a complication for each individual patient. This figure was attributed without knowledge of the post operative outcome for the patients evaluated (blinded attribution).

The patient records and electronic coding of post operative complications at 90 days were assessed and compared against the risk ratio percentage attributed by the model. Complications were defined as: an extended length of stay in hospital beyond the expected average, the occurrence of postoperative complications, encompassing a wide range of medical conditions or mortality (Table 2).

At Northumbria, stratification into low vs high/moderate risk was also compared to a consultant (attending) anaesthetists (anaesthesiologists) and preoperative assessment teams, allowing comparison between the AI model stratification, and current consultant led care.

## Results

445 patients were assigned a risk ratio by the AI model, 302 patients in Swansea, and 143 at Northumbria. Patients were categorised either into a high/moderate, or a low-risk group. In total, the model identified 192 high/moderate risk patients with a risk ratio greater than 6%, and 253 low risk patients with a risk ratio of less than 6%.

There were variations in the distribution of risk at the two sites (Table 3).

**Table 3:** Percentage of patients in each risk group; Swansea Bay vs Northumbria Healthcare.

|  | Swansea | Northumbria |
| --- | --- | --- |
| **Low Risk** | 52% | 32% |
| **High/ Moderate** | 48% | 68% |

A total of 37 patients from both sites had post operative complications. 26 patients (from the 37 who did have a complication) were correctly identified by the AI model as “high risk”. Therefore 11 patients (who did have a complication) were incorrectly identified by the AI model as “low-risk”. This gave a sensitivity of the AI model to correctly predict the risk category (indicated by complications) of 70%.

The Negative Predictive Value (NPV), an indication of how well the model identified those patients who were not high risk and did not have a post operative complication was 96%.

Overall complications are shown (Table 4) and further analysed in the results matrix (Table 5).

**Table 4:** Complications by AI Risk Category.

|  | No Complication | Complication | Totals |
| --- | --- | --- | --- |
| **Low Risk** | 242 | 11 | 253 |
| **High/Moderate Risk** | 166 | 26 | 192 |
| **Totals** | 408 | 37 | 445 |

Negative Predictive Value (NPV) 96%
Sensitivity 70%

**Table 5:** Consultant(Attending) led care risk prediction in Northumbria.

|  | No Complication | Complication | Totals |
| --- | --- | --- | --- |
| **Low Risk** | 102 | 3 | 105 |
| **High/Moderate Risk** | 31 | 7 | 38 |
| **Totals** | 133 | 10 | 143 |

There was one death in a patient who was identified as high-risk with a risk score of 34.4%. The model performance was then compared to consultant (Attending) led-care standard care for a subset of 143 Northumbria patients. Each patient was designated into high/moderate vs low risk by a consultant anaesthetist. The categorisations between Consultant and the AI model were descriptively compared (Table 5 & Table 6). There were negligible differences in sensitivity, with consultant led care having a sensitivity of 70% and 70% for the AI model. The Negative Predictive Value was marginally higher at 97% compared to 96% respectively. In comparing the models’ outputs to consultant led care, the model demonstrated a high degree of equivalence in matching the passement opinion (Table 6). The model match 84% (n=32) of those patients deemed high or moderately high risk by preassessment teams (n=38) and 92% of those considered as lower risk and suitable for high volume low complexity pathways.

**Table 6.** comparing consultant (attending) review and identification of risk category to AI prediction in the Northumbria cohort.

| N=143 | True risk category<br>(according to<br>complication event) | Consultant Led<br>Care (CLC)<br>Categorisation | AI Model<br>Categorisation in<br>agreement to CLC |
| --- | --- | --- | --- |
| **Complication in**<br>**high-risk group** | 7 | 38 | 32 (84%) |
| **Complications in**<br>**low-risk group** | 3 | 105 | 97 (92%) |
| **Model Sensitivity** |  |  | 71% |
| **Model Specificity** |  |  | 68% |
| **Model NPV** |  |  | 97.1% (92.9% 98.8%) |

Complications were further analysed with a Clavien -Dindo index^21^ to review the severity of complications for high, moderate, and low risk patients (Table 7). The Clavien-Dindo classification is a system for grading the severity of postoperative complications (PPCs) and is used as a tool for assessing and reporting PPCs in surgery. The classification is based on the type of therapy needed to correct the complication. Complications are graded from mild requiring minor intervention for example (I), infections requiring antibiotics (II), further surgical procedure (IIIb), and death (V). More complex complications generally were seen in higher risk patients.

**Table 7:** Outcome of risk classification compared with severity score based on Clavien-Dindo (CD) classification.

|  | Severity (CD Classification) |  |  |  |  |  |
| --- | --- | --- | --- | --- | --- | --- |
|  | CD I | CD II | CD IIIa | CD IIIb | CD IV | CD V |
| **Low** | 8 | 2 |  | 1 |  |  |
| **Moderate** | 2 | 3 | 2 |  |  |  |
| **High** | 6 | 6 | 1 | 2 |  | 4 |

**Table 8:**
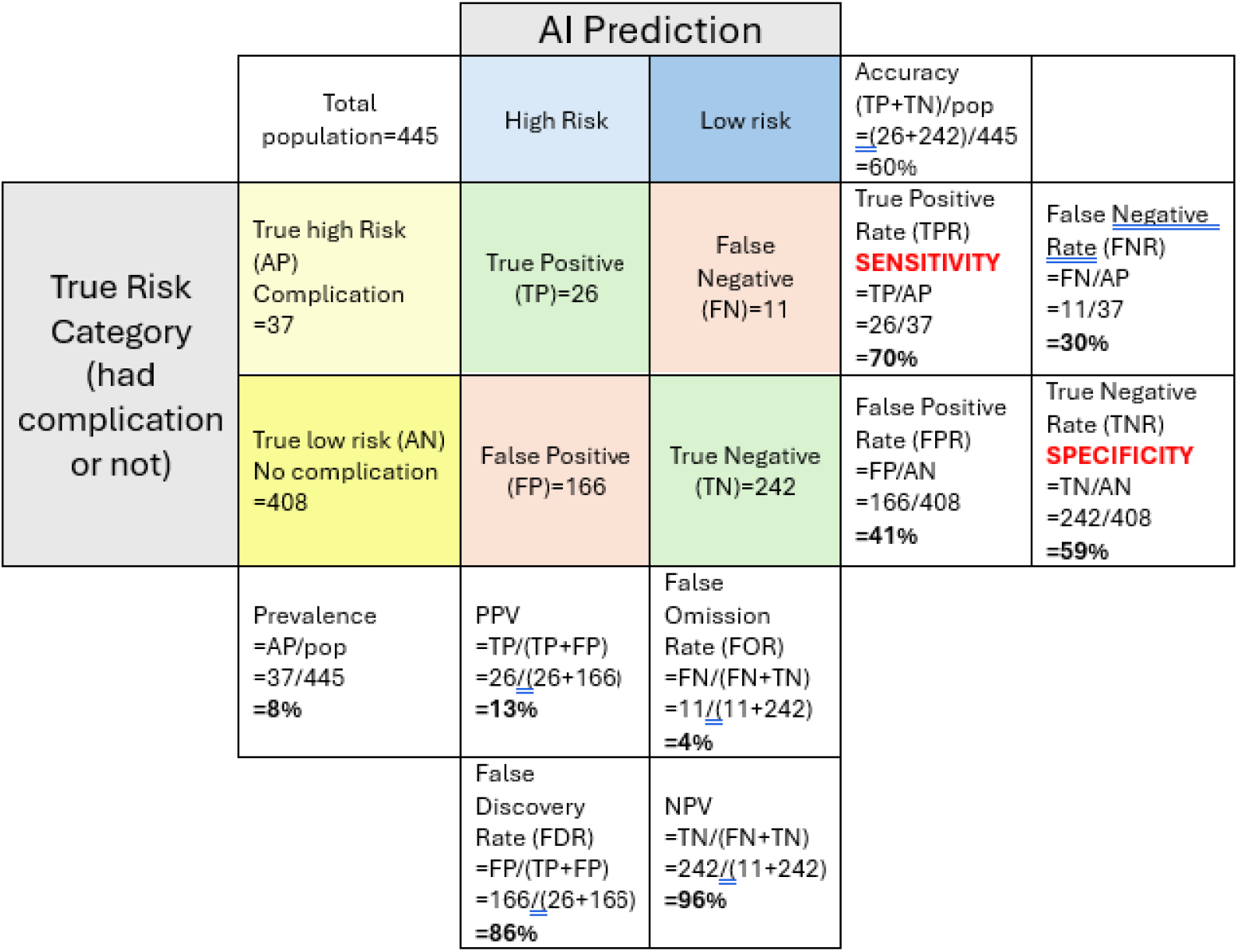
Matrix of AI Prediction Results vs High and Low Risk.

## Discussion

Growing orthopaedic waiting lists in the UK, especially in the years following the COVID 19 pandemic ^1,2^, and the increased use of ambulatory surgery centres in the United States, pose a huge challenge for perioperative teams assessing risk. The model devised provides a machine learning solution designed to act as a clinical decision aid to support the stratification and prioritisation. The polynomial regression model developed uses a statistical technique commonly employed in risk stratification. This work describes the creation of a model based on risk ratio thresholds drawn from a large population (n=6,730). It is important to note that the risk ratio threshold chosen for a given population or healthcare system could be varied according to the local site-specific hospital services provided in terms of directing patients appropriately according to the level of care.

When used to assess the risk for adverse outcomes following hip and knee replacement surgery, it allows accounting for nonlinear relationships between patient characteristics and surgical outcomes. It is more complex than simpler models such as logistic regression, but less complex than a neural network, which may become more over reliant on training data. It provides a useful middle ground for predicting surgical risk. ^22,23^ This validation study supports its potential use in stratifying risk in primary hip and knee replacement. Risk stratification performance into High/Moderate groups was adequate and similar to consultant lead care, with overall categorisation of patients suitable for high volume low complexity sites also equivalent.

The validation exercise was driven from identifying individuals with a high risk of complications and the potential need of higher level care. There was an 8% prevalence of true complications in the total population cohort. The sensitivity of the model in predicting correct classification (high risk), and using post operation complications as the marker, was 70%. This relatively high sensitivity provides reassurance that most patients with likely complications can be predicted, and the sensitivity value compares equivalently with that for current consultant/Attending led assessment. Note, it does leave a false negative rate of 30% - meaning that 30% of patients predicted to be low risk (not have a complication) may be at risk of developing a complication, and would be considered as higher risk. However, once again this compares with current standard of consultant led preoperative assessment review and screening.

It is acknowledged that perfect complication prediction is near impossible, and the AI performs at least as well as current screening processes. Importantly, the negative predictive value (NPV) of having a listed complication was 96%. This supports the notion that, if deemed low risk by the model, a patient is less likely to experience a complication. Note the likelihood of having no complications in this population was 92%. The high negative predictive value indicates the model’s performance in correctly identifying patients who require less intervention postoperatively. In the context of HVLC it is important that patients that are directed to elective hubs and ASC have high probability of having routine, uncomplicated recovery, without the need to full preoperative assessments.

The results do show a substantial number of false positives for high-risk categorisation (n=166/408). This 41% false positive rate means that many patients will be classified by the AI model as high risk, despite having complication free procedures. If used alone, it could result in many patients being referred or directed to high level care facilities for their operation, somewhat unnecessarily. However, the caveat is that from a safety perspective this is a more desirable situation than patients with higher needs being referred to sites without adequate post operative emergency care.

One patient died in the post-operative period. They were identified as being very high risk, with a risk score of 34.4%. The model potentially has an ability to identify the most high-risk patients, though this should be evaluated further with a larger number of patients. This aspect of the model is useful in being able to select such patients earlier for consultant lead pre assessment and potential preoperative health maximisation. It also highlights the usefulness in providing an individual risk ratio for complications that may allow a serious discussion with a patient about their surgical risks, and perhaps on occasion whether to proceed with elective surgery at all.

Clinically the findings and use of the model need to be seen in context of the speciality and procedure, and this may not be the same for other specialties and operations. The prevalence rate for complications in hip and knee replacement (orthopaedic surgery) is very low. This study showed a prevalence of 8%. This low prevalence means that the default allocation for patients should be to a HVLC site. It is, however, important to screen and identify those relatively few patients who are at high risk of complications. The AI model can identify such patients in 70% of cases at an early stage. In turn this also could reduce the burden on pre assessment, by potentially identifying lower risk patients for a lighter, or streamlined, preoperative assessment pathway. Such patients could be triaged early to elective HVLC surgical hubs or ASC with potential savings in staffing and improved pathway efficiencies.

Waiting list risk ratios could assist efficient planning of resources, with the ability to forward plan list numbers at different hospital sites, with an understanding and planning for anaesthetic complexity, together with high dependency or critical care resources that may be required.

Complications are often severe and may require additional medical intervention or long-term hospital stays. Some of the complications within the target variables are well known and potentially life-threatening, such as stroke, myocardial infarction, pulmonary embolism, and deep vein thrombosis. Other complications, such as urinary retention or ileus, may be less serious, but still have a significant impact on the recovery of the patient.

While a machine-learning tool offers a sophisticated method in planning preoperative assessment, it will have limitations and cannot be devoid of potential error. No risk predictor can be perfect in identifying patients that will go on to have complications and it must be remembered that all patients are at risk. Indeed, in the study there were complications in patients predicted as low risk. Many of these complications would have been difficult to predict during the standard pre-operative review. For example, urinary retention, in a well patient, with no history of benign prostatic hypertrophy. However, data points not considered by the model may have highlighted these somewhat; for instance, a wound infection in patient with a high body mass index, which is a known risk factor for wound infection. Additional work may enhance further the accuracy of the model, for example looking at a larger group of patients. However, the current study suggests that the model was at least as accurate as consultant / attending led preoperative assessment in identifying high and low risk patients.

It is important to note that the accuracy of a machine-learning tool is dependent on the data available and inputted from patient records. This must be up to date, secure, and accurate. This highlights that the use of a machine learning model such as this should be as a useful efficient clinical adjunct, with targeted clinician oversight, and cannot be employed in isolation. The model and AI capability will improve with time as more data is incorporated.

The benefit level of AI depends on the acceptability of staff and patients, its application and evaluating if combining ML with human clinicians will outperform the care provided by either system alone. We believe that this study supports the use of validated machine learning models in shared decision-making by informing healthcare professionals about the risk of complications, thus aiding in the management of elective surgery backlogs and changing trends in perioperative care in Ambulatory Surgery Centres.

## Conclusion / Summary

An artificial intelligence (AI) model was developed for risk stratification in hip and knee arthroplasty. The model was capable of assigning risk scores for postoperative complications and could categorise patients into high and low risk. A validation study in 445 patients showed the AI model had a sensitivity of 70% for correctly predicting high risk and negative predictive value of 96%. This AI risk prediction performance was comparable to consultant-led standard review and risk stratification. AI has the potential to support more streamlined and efficient preoperative risk stratification, potentially reducing the burden on preoperative assessment teams and optimising resource allocation offering a solution to elective surgery backlogs.

## Data Availability

All data produced in the present work are contained in the manuscript

https://github.com/fabrylab/MP_ventilation

## ACKNOWLEDGEMENTS

This study was partly funded by a Welsh Assembly Government Transformation Grant towards the OWL-I (Orthopaedic Waiting List Initiative) at the Swansea University Health Board.

We are grateful for the support provided by the Digital Informatics and Information Technology departments in Swansea and Northumbria, as well as the waiting list management teams in both organisations

